# A Stacking Framework for Polygenic Risk Prediction in Admixed Individuals

**DOI:** 10.1101/2024.01.31.24302103

**Authors:** Kevin Liao, Sebastian Zöllner

## Abstract

Polygenic risk scores (PRS) are summaries of an individual’s personalized genetic risk for a trait or disease. However, PRS often perform poorly for phenotype prediction when the ancestry of the target population does not match the population in which GWAS effect sizes were estimated. For many populations this can be addressed by performing GWAS in the target population. However, admixed individuals (whose genomes can be traced to multiple ancestral populations) lie on an ancestry continuum and are not easily represented as a discrete population.

Here, we propose slaPRS (**s**tacking **l**ocal **a**ncestry **PRS**), which incorporates multiple ancestry GWAS to alleviate the ancestry dependence of PRS in admixed samples. slaPRS uses ensemble learning (stacking) to combine local population specific PRS in regions across the genome. We compare slaPRS to single population PRS and a method that combines single population PRS globally. In simulations, slaPRS outperformed existing approaches and reduced the ancestry dependence of PRS in African Americans. In lipid traits from African British individuals (UK Biobank), slaPRS again improved on single population PRS while performing comparably to the globally combined PRS. slaPRS provides a data-driven and flexible framework to incorporate multiple population-specific GWAS and local ancestry in samples of admixed ancestry.

## 1.2 Introduction

Since the first genome wide association study (GWAS) published in 2005, GWAS have successfully implicated thousands of risk variants across a variety of traits^1^. While a single risk variant may only explain a small percent of a trait’s heritability, a sizable proportion of phenotypic variation can be explained by summarizing an individual’s genetic risk for a given disease or trait in polygenic risk scores^2^ (PRS). PRS are typically computed as a weighted sum of risk alleles using estimated effects from an external GWAS as weights. These PRS have been used^3,4^ to identify individuals at high risk of disease, improve diagnostic accuracy, and allow for tailored personalized treatment for disease risk prediction in complex traits including coronary artery disease^5,6^, type 1 and 2 diabetes^7,8^, breast cancer^9,10^, and more^11^. However, PRS fail to capture the full variability expected from heritability estimates while also being susceptible to environmental confounding and indirect genetic effects such as assortative mating^12–14^.

Furthermore, performance of a PRS in predicting a phenotype for a target sample can be ancestry dependent. In particular, PRS prediction performance decays as genetic divergence increases between the target sample of interest and external GWAS^15,16^. This performance decay can mainly be attributable to 1) differences in allele frequencies and 2) differences in both marginal and causal effect sizes of variants across populations^17^. Causal effect sizes themselves can differ across populations due to unique environments and demography, though recent work in admixed individuals has suggested causal effect sizes are shared across populations^18^. However, even when causal effects are shared, marginal estimated GWAS effect sizes can still differ due to differences in linkage disequilibrium (LD) tagging the true causal variant. The extent in how LD differs across populations varies along the genome^19^, prompting work in the transferability of PRS across diverse populations to often consider a local approach in combining genetic evidence^20,21^. Specifically, approaches often model local population-specific LD patterns in regions to better identify true local risk variants and increase effective sample size^21^.

The ancestry dependence of PRS is further exacerbated in the context of admixed individuals. Historically, genetic studies group admixed individuals of varying ancestry proportions into a single discrete ancestral label such as “African American” or “Hispanic”. However, the genetic ancestry in an admixed sample varies across both individuals and regions prompting a recent push to consider ancestry on a continuum rather than as discrete ancestral groups^22^. In admixed individuals, Bitarello and Mathieson showed predictive accuracy of a PRS for height using European summary statistics increased linearly with global European ancestry proportion across various datasets^23^. Similarly, Cavazos and Witte showed in simulations a similar linear relationship with both European and African summary statistics performing better as the proportion of European and African ancestry respectively increased across admixed samples^24^. Such ancestry dependence of PRS in admixed individuals is problematic even if all ancestral groups have predictive PRS, as admixed individuals that have most of their genetic ancestry from one parental group will benefit more from potential downstream clinical utility of PRS than groups with equal contribution from both ancestries. Even developing PRS specifically for the admixed group will not ameliorate this problem as such a PRS will only work well for admixed individuals with admixture proportions similar to the group mean. While the field of genetics has acknowledged and begun making strides in addressing inequity in genomic research^25,26^, development of methods to construct well-performing PRS free of ancestry dependence in admixed samples is needed.

To overcome the ancestry dependence of PRS performance using a single population GWAS in admixed samples, recent work has proposed methods that leverage GWAS summary statistics from the multiple ancestral populations of an admixed sample. Incorporating GWAS effect sizes from multiple populations provides many benefits, including identifying population specific risk variants and boosting sample size if risk variants are shared. In admixed African Americans, methods have been proposed that 1) consider local ancestry by matching chosen risk variants with an individual’s local ancestry at that position^23,27^ and 2) ignore local ancestry and construct a joint PRS as a linear combination of global European and African PRS^28^. In simulations, Cavazos and Witte conducted a comprehensive review of both approaches^24^. While the first approach, deconvoluting ancestry and matching risk variants on population-specific GWAS effect sizes, was initially suggested to perform well^27^, this result failed to consistently replicate as shown in Cavazos’ simulations and Bitarello’s real data application^24,27,28^. Surprisingly, the second approach ignoring local ancestry information (linear combination of global European and African PRS) was found to efficiently optimize prediction across a range of European ancestry quantiles in admixed African American individuals. However, use of global population specific PRS ignores the unique local admixture present in any given region within a sample of admixed individuals, missing potential population specific risk variants in a region or local GxG interactions on a specific ancestral background. Thus, it is possible that performance of local population specific PRS (i.e., a PRS using only risk variants in a genomic region and a specific population GWAS effect sizes) will vary across admixed individuals.

In this work we propose slaPRS (**s**tacking **l**ocal **a**ncestry **PRS**), a novel stacking framework to construct admixed PRS for quantitative traits that combines local population specific PRS constructed using population specific effect sizes in local genomic regions. Stacking is an ensemble machine learning method that aims to optimize prediction accuracy by combining separate prediction models^29,30^. In target samples of a single ancestry, Prive et al successfully used stacking to optimize the commonly used clumping and thresholding (C+T) PRS method through deriving a linear combination of PRS across all possible parameters, rather than learning a single set of optimal parameters^31^. Outside of PRS construction, stacking has been used in other genetic methods such as the recent REGENIE method for GWAS that improved computational efficiency through orders of magnitude by conditioning on the predicted individual trait values from combining local polygenic risk predictors^32^. In our approach, we first divide the genome into windows of a predetermined size and in each local window compute population specific local PRS using the respective population specific GWAS effect sizes via C+T. In training data, we then fit a penalized regression model to combine local population specific PRS across the genome to determine unique weights that are used to predict the phenotype in testing data. We show in extensive simulations and real data application of admixed African Americans and African British that slaPRS removes the ancestry dependence of PRS performance present in traditional single-population GWAS PRS and outperforms or compares similarly to existing methods in an efficient data-driven process.

## 1.3 Methods

Consider a sample of N admixed individuals with ancestral contributions from population A and B (slaPRS is not restricted to two-way genetic admixture but is assumed here for notational simplicity). Let **X** be the *NxM* admixed genotype matrix (M is the total number of variants genome wide) and **Y** the *Nx*1phenotype vector. Let *L*_ij_ be an *NxM* matrix denoting the haplotype-level local ancestry (*l*_ijl_, *l*_ij2_.) of individual *i* at marker *j* We assume the phenotype can be expressed as:

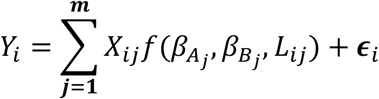

Where *X*_ij_ is the genotype dosage for individual *i* at marker *j*, and 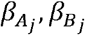 are effects for marker *j* on the phenotype in populations A and B respectively. Here, 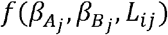 is a weighted average of population specific GWAS effect sizes and local ancestry (see supplementary for derivation):

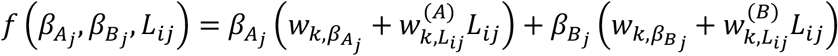

Where 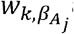 and 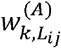 (and similarly for population B) are weights for population A effect sizes 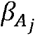 and local ancestry interaction in each genomic region *k* that are learned via ensemble learning (stacking) in the slaPRS framework (see details below).

### 1.3.1 slaPRS Framework

We developed slaPRS for constructing admixed PRS using three main features: 1) a local window approach 2) local population specific PRS and 3) an ensemble stacking framework to combine local population specific PRS. For slaPRS, we assume existence of GWAS effect size estimates for each ancestral population in an admixed population. We first partition the admixed genotype matrix into K non-overlapping genotype blocks *G* = {*G*_1_,*G*_2_,…*G*_K_} with blocks predefined by physical distance. In our analysis we considered blocks spanning 1Mb and 5Mb of physical distance, each with *m*_*k*_ SNPs such that 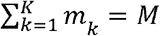.

#### Level 0 Local Population-Specific PRS and Ancestry

In the training set of admixed individuals, in each block *G*_*k*_ across the genome (using the *m*_*k*_ SNPs in the block) we first separately computed vectors of local population A PRS (*A*_*k*_)and local population B PRS (*B*_*k*_) using clumping and thresholding (C+T). While C+T was used in slaPRS, any PRS construction method could be used in our framework. In this step, each block’s C+T optimized ancestry PRS can be viewed as a level 0 model prediction to be stacked in our stacking framework (Figure 1). Clumping first removes variants in strong LD with others using in-sample LD for that region, while greedily retaining the most significant variants^33^. Varying p-value thresholds *p =* {5*e*−2,5*e−*4,5*e*−6, 5*e*−8} were considered (cross validation in Level 1 stacking model used to select optimal to use in testing set) to construct ancestry-specific local PRS in each block using the respective population’s estimated effect sizes. In this step, we make no assumption on whether risk variants are shared across ancestral populations, and thus local PRS *A*_*k*_ and *B*_*k*_ can have varying risk variants.

**Figure 1.**
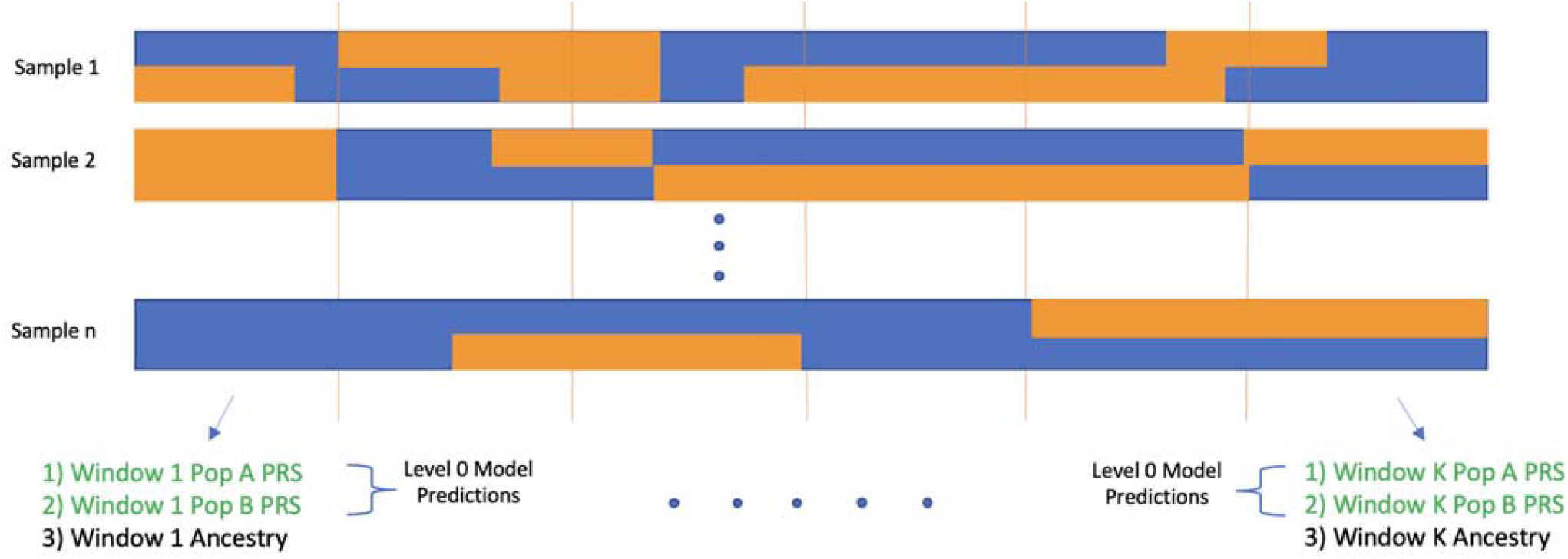
Diagram of local window and level 0 population specific PRS model predictions. Admixed genomes split into 5Mb windows and in each window a local population A and B PRS are computed using population-specific effect sizes. Local ancestry further computed to form covariate vector for level 1 stacking model.

For each sample, we computed the *Nx*1 vector of local ancestries *Anc*_*k*_ in block as the % of population A ancestry. We constructed interaction terms *A*_*k*_ * *Anc*_*k*_ and *B*_*k*_ * *Anc*_*k*_ to allow for the effect of the local population *A*_*k*_ PRS *B*_*k*_ and to vary by a given ancestry. Following completion of level 0 in our framework, block *k* has the covariates (Figure 1):

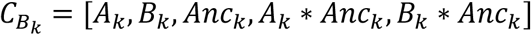

After aggregating the B total local block covariates across the genome, let C be the *N x* (*k x* 5) matrix:

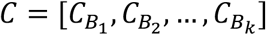

#### Level 1 Elastic Net Stacking Model

We then trained an elastic net^34^ penalized regression model to stack the local level 0 predictions (local population-specific PRS and ancestry) across the genome. The population’s GWAS that optimizes the local PRS can vary across the genome (see introduction) in an admixed sample, and stacking provides a data driven approach to inform which population’s local PRS should be upweighted or shrunk. We used elastic net, which combines ridge regression^35^ and LASSO^36^, because the genetic architecture of a trait is unknown a priori (unknown which local blocks harbor causal risk variants and the distribution of local block heritability). When most local windows are weakly informative, ridge tends to have higher prediction accuracy while LASSO would likely outperform when only a small number of local windows are highly informative. Elastic net allows a data-adaptive approach to inform the amount of shrinkage and whether shrinkage patterns should favor ridge or LASSO to best accommodate a trait’s genetic architecture.

To determine which aspects of our stacking framework drives increases in PRS performance, we considered three level 1 elastic net stacking models that vary in the covariates included from block *B*_*k*_:

1. Local population A PRS only

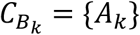
2. Local population A and B PRS only

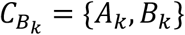
3. Local population A and B PRS, Ancestry and Interactions

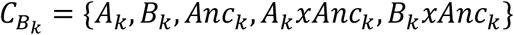

Model 1 considered only local population A PRS *A*_*k*_ to investigate how stacking local PRS alone improves compared to a global population A PRS. Model 2 added local population B PRS *B*_*k*_ to assess the benefit of adding population B GWAS information, while Model 3 further included ancestry and interaction terms to allow for the effect of a local population specific PRS to vary based on ancestral background. Total covariates in each proposed level 1 model aggregate covariates 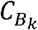 across all blocks genome wide.

For each considered model, we fit a level 1 elastic net model^34^ to combine the level 0 ancestry-specific PRS and additional covariates across the genome.

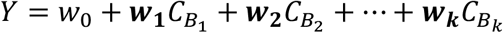

Where ***w***_**1**_,***w***_**2**_,**…*w***_***k***_ are vectors of regression coefficients from the covariates in 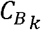 Estimates of ***w***_***k***_ in the above model given the genome wide covariate matrix are obtained by minimizing the penalized objective function with respect to *β*:

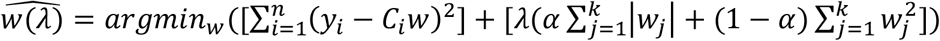

Parameter *λ*determines the amount of shrinkage in model coefficients while *α ∈* [0,1] balances the L1 and L2 penalty from ridge regression (*α* = 0)and LASSO (*α* = 1)To optimize all parameters including the p-value threshold *p* = {5*e* – 4,5*e* – 6,5*e* – 8} used in constructing level 0 local ancestry PRS via C+T, *α =* {0,0.1,0.2,…,1} and *λ*={10^−3^,…,10^3^}, we employed K-fold cross validation with 10 folds and selected the set of *p, α λ*, and that produced the lowest adjusted *R*^2^.

Estimates of ***w***_***k***_ for each block across the genome can be used (see supplementary for derivation) to express the weight for each variant in PRS construction to be a linear combination of population 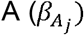 and B 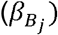 GWAS effect sizes and learned block weights:

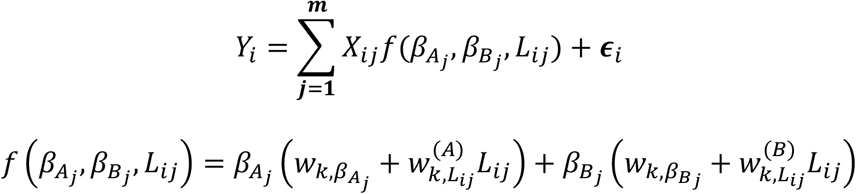

Where 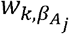 and 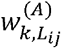 (and similarly for population B) are weights for population A specific local PRS *A*_*k*_ and its local ancestry interaction term.

Once weights from the level 1 elastic net stacking models had been estimated from the training data, in testing data we then computed the same level 0 model predictions and covariates in each block and aggregated genome wide:

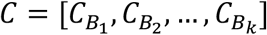

Where 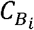 is defined as one of the three considered level 1 models. We then predicted trait values using estimated weights from the elastic net model:

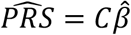

The estimated PRS is then tested against simulated phenotypes or trait values in real data.

#### Genotype, Phenotype, and Population-Specific GWAS Simulation

For our simulations and real data applications we focused on admixed African Americans/British with European and African ancestral backgrounds. To simulate genotype and phenotype data for an African and European population with realistic allele frequencies and linkage disequilibrium patterns, we used the coalescent-based pipeline as described by Martin et al^16^ and Cavazos et al^16,24^. Using msprime^37^ with an out-of-Africa demographic mode modeling HapMap^38^ chromosome 20 haplotypes, we simulated n=10,000 European samples and varying African sample sizes n={2000, 5000, 10,000}. Simulated population specific genotypes were then used to estimate marginal variant effect sizes.

We then simulated quantitative trait phenotypes using the simulated genotypes. We first assumed complete transethnic sharing of genetic architecture across African and European populations, in which true causal variants, causal effect sizes, and overall heritability are consistent across populations. Under this scenario, performance of estimated PRS should vary only because of differences in allele frequency and LD across population. We subset variants with minor allele frequency > 5% in both populations and randomly sampled m={100, 500} shared causal variants. True causal effect sizes were drawn from a normal distribution 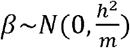 where *h*^2^ = {0.10,0.30} is the SNP-based heritability. In results, we focused on the most realistic simulation scenario consisting of *h*^2^ =0.10 and *m* =100. We then considered the simulation scenario in which genetic architecture differs across ancestral populations by assuming true causal variant locations and overall heritability are shared, but now simulating causal effects 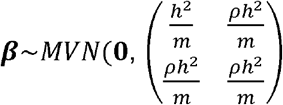 varying transethnic genetic correlation ρ = {0.20,0.50,0.80}.

In both simulation scenarios, the true genetic score *G* was then defined as the product of sampled causal genotypes and their respective simulated effect sizes 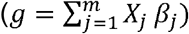, standardized to ensure total heritability of 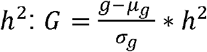. We then simulated the environmental effect from a normal distribution with variance comprising the remaining phenotype variance ∈ ∼ *N* (0,1 – *h*^2^)and similarly standardized: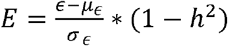. We defined phenotype data Y for both populations as the sum of the standardized true genetic score and using a linear model environmental effect *Y* = *G* + *E*. We then estimated effect sizes 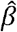 for each variant genome wide using a linear model Y = XB + ∈, using each population’s respective simulated phenotype and genotype data.

We additionally simulated n=1,000 European and n=1,000 African founder samples to simulate n=10,000 admixed African Americans genotypes via RFMix^39^ with s=12 generations of admixture for training and testing slaPRS. Simulated admixed genotypes had known phase and known local ancestry. We followed the same pipeline described above to generate the phenotype given the simulated genotypes. In the scenario where causal effects differed across populations, we considered haploid chromosomes *H*_ij1_ and *H*_ij2_ (corresponding haplotype 1 and 2 for individual *I* at variant *j*) and matched the population specific effect sizes on the local ancestry of a variant’s haplotype background to derive the true genetic component: 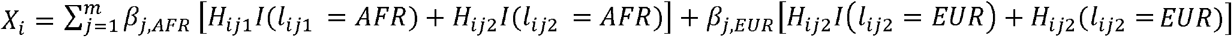. To prevent issues of overfitting, we split our sample into testing and training data using a 70:30 split, resulting in n=7000 and n=3000 admixed samples in the training and testing data splits. The outlined simulation procedure was repeated 150 times to evaluate slaPRS and perform method comparisons.

### 1.3.2 Comparison of Methods

#### Clumping and Thresholding (C+T)

We first compared the proposed slaPRS method against global single population PRS, *PRS*_*EUR*_ and *PRS*_*AFR*_, constructed using clumping and thresholding (C+T) with GWAS effect sizes from the respective population separately. In the C+T algorithm, we first clumped SNPs using each population’s GWAS effect sizes with a window size of 250Kb and linkage threshold *r*^2^=0.10 and then optimized the threshold parameter in the 70% training set with - log_10_(*p*) p value thresholds including {1, 2, …, 8}. The threshold that optimized PRS performance was then used in the 30% testing set to retain clumped risk variants to include in the PRS construction.

#### Linear Combination of Global Population Specific PRS

The second approach compared against was the method proposed by Marquez-Luna et al^28^ which constructed a PRS as a linear combination of two global population-specific PRS:

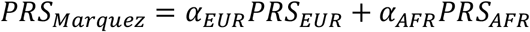

Here, *PRS*_*EUR*_ and *PRS*_*AFR*_ are the same global PRS constructed using C+T and the respective population GWAS as described above. To estimate the mixing weights (α_*EUR*_, α_*AFR*_ )and global polygenic risk scores (*PRS*_*EUR*_, *PRS*_*AFR*_), we followed proposed guidelines and used cross validation. The 70% training set of admixed samples was first split in half, where the first half was used to estimate the thresholding parameter in the C+T algorithm. In the second half we constructed *PRS*_*EUR*_ and *PRS*_*AFR*_ using the optimal p-value threshold from the European GWAS (as is typically larger), as done by Marquez-Luna et al. In this same second half of the training set, we then estimated *α*_*EUR*_ and α_*AFR*_ by finding the least squares estimates to:

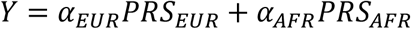

With the optimal p-value threshold and mixing weights *α*_*EUR*_ and α_*AFR*_ derived from training data, we then constructed *PRS*_*Marquez*_ as the weighted sum of *PRS*_*EUR*_ and *PRS*_*AFR*_.

### 1.3.3 Quantifying Performance of Estimated PRS

To quantify and compare performance of each PRS across methods, we computed the proportion of variance explained (adjusted *R*^2^) of the simulated quantitative phenotype with the To quantify and compare performance of each PRS across methods, we computed the estimated PRS adjusting for % European ancestry. Because one of our main objectives is to create a PRS with performance independent of the global ancestry of an admixed individual, we further stratified our adjusted *R*^2^performance metric by European ancestry quantiles [0-20%, 20-40%, 40-60% and 60-80%, 80-100%]. We also compared the mean simulated phenotype value in the top 10% PRS quantile with the bottom 10% PRS quantile to assess the PRS’ ability to identify high-risk and low-risk individuals.

### 1.3.4 Real Data Application

We evaluated slaPRS in real data applications using n=20,262 admixed African British individuals in the UK Biobank^40^. To choose samples, we selected admixed samples falling on the diagonal between the European and African corners of the PC plot (Supplementary Figure 1). We used autosomal imputed genotypes in constructing polygenic risk scores. Phenotype data included the lipid biomarkers LDL, HDL, and total cholesterol. Lipid biomarker phenotypes were chosen because the Global Lipids Genetic Consortium^41^ had collected large sample (excluding UK Biobank samples) ancestry specific GWAS in Europeans (n=1.32 million) and Admixed African or Africans (N=99.4k). For all 20,262 samples we inferred local ancestry with genotypes first phased using BEAGLE 5.0^42^. We used RFMix^39^ to infer local ancestry using phased haplotypes from European and African subpopulations from 1000 Genomes^43^ individuals as references. From inferred local ancestry, we further computed global ancestry using tract lengths for sample stratification. We split the admixed dataset into 70% training and 30% testing for model training and method comparison.

Because the true PRS is unknown in real data, to quantify PRS performance across methods we computed the proportion of variance explained (adjusted *R*^2^) between the estimated PRS and phenotypic value (instead of true genetic score) from the model including the first 4 principal components:

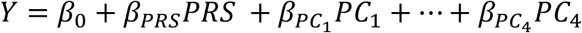

Similar to simulations, we computed adjusted *R*^2^ across the entire testing sample and then also stratified by European ancestry quantiles. We also compared the mean simulated phenotype value in the top 10% PRS quantile with the bottom 10% PRS quantile. Performance metrics were computed with the median reported over 50 folds.

## 1.4 Results

### 1.4.1 Comparison of PRS Performance Assuming Shared Genetic Architecture across Ancestral Populations

To evaluate the performance of slaPRS, we first conducted simulations with complete sharing of genetic architecture across ancestral populations (i.e., true effect sizes and risk variants are shared across European and African populations) for various disease architectures (see methods). Under this setup, differences in GWAS estimated effect sizes across ancestral populations are a function of solely LD. We constructed our stacked PRS using simulated European and African GWAS effect sizes for simulated admixed African Americans of varying ancestry proportions. The distribution of overall European ancestry in our simulated admixed African Americans was approximately normally distributed with a mean of around 50% (Supplementary Figure 2).

We focus first on the full level 1 model with 5Mb windows using the local African and European PRS and local ancestry information in each block 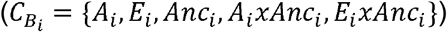 with heritability *h*^2^ = 0.10, number of causal variants *m* = 100, and equal size European and African GWAS sample size *n* = 10,000. Across simulations, our stacked PRS generally had an increased adjusted *R*^2^ with the simulated phenotype compared to the existing approaches. slaPRS had a 5.93% median adjusted *R*^2^for the true PRS across all admixed individuals in the testing set compared to C+T *PRS*_*EUR*_ (3.17%) and *PRS*_*AFR*_ (3.18) and *PRS*_*Marquez*_ (3.39%) that globally combines *PRS*_*EUR*_ and *PRS*_*AFR*_. Comparing individuals in the top vs bottom 10% of the PRS distribution, slaPRS had higher trait stratification ability with larger mean differences (0.84 vs 0.62, 0.64, 0.64 for *PRS*_*EUR*_, *PRS*_*AFR*_ and *PRS*_*Marquez*_ respectively). We further stratified testing samples by quantiles of European ancestry and found our stacking approach using the full model explained more variance of the phenotype compared to both *PRS*_*EUR*_, *PRS*_*AFR*_ and *PRS*_*Marquez*_. Across all ancestry quantiles the percent increase in median adjusted *R*^2^ for slaPRS compared to the other methods ranged from 38.46% to 120.61% (Figure 2). Most notably, slaPRS strongly reduced the ancestry dependence of PRS performance as compared to *PRS*_*EUR*_ and *PRS*_*AFR*_. When quantified through a simple linear model, the adjusted *R*^2^ for slaPRS increased by 0.0009 for every European ancestry quantile increase ranging from 5.69% (0-20% European ancestry) to 5.91% (80-100% European ancestry). On the other hand, single population *PRS*_*EUR*_ and *PRS*_*AFR*_ had larger changes in *R*^2^ of 0.004 (2.60% to 4.22 %) and -0.001 (4.11%-3.60%) respectively for every quantile increase. *PRS*_*Marquez*_ compared similarly to slaPRS with an increase of 0.0008 for every quantile increase, ranging from 3.46% to 3.91%.

**Figure 2.**
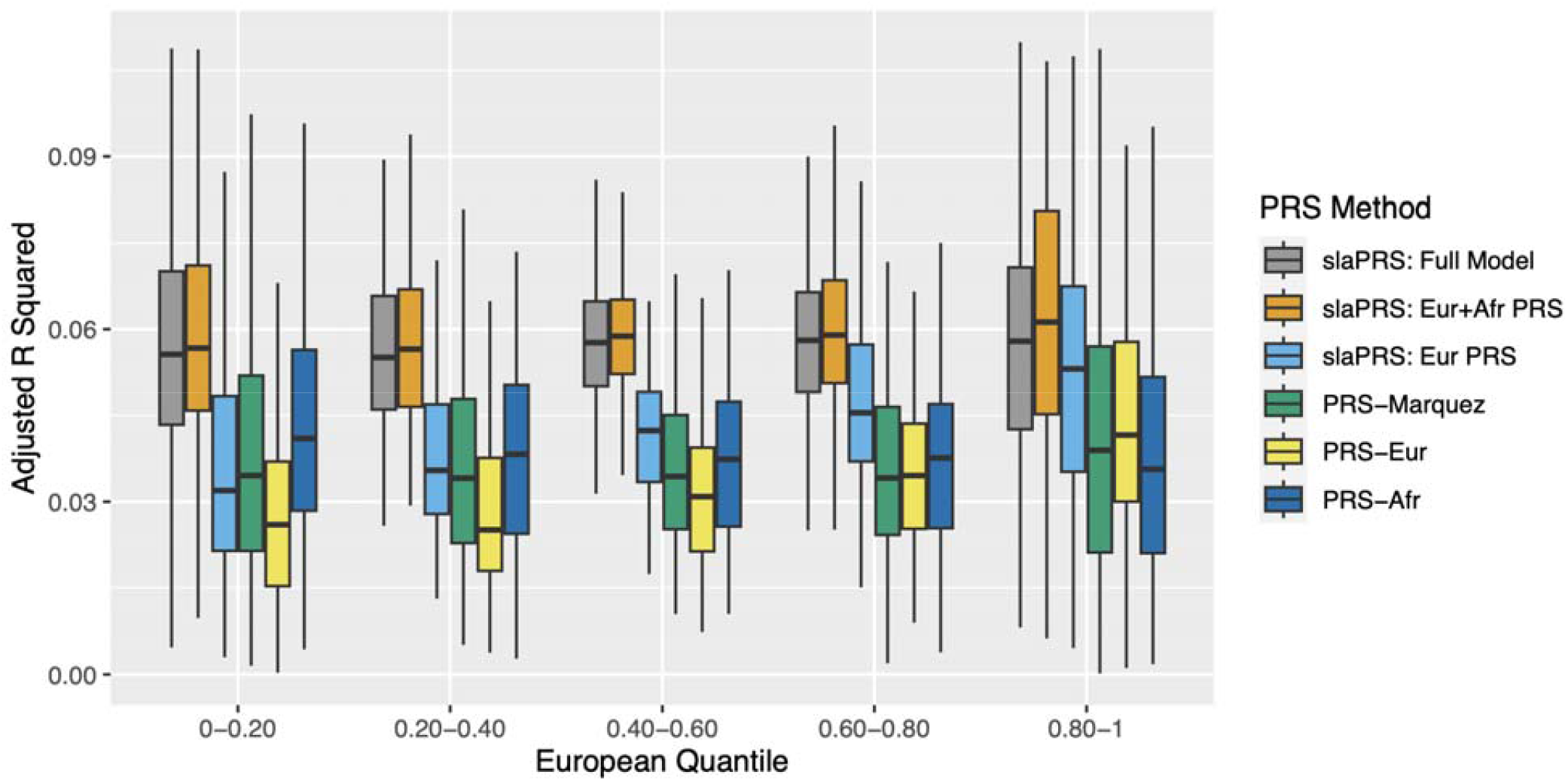
Error! No text of specified style in document. 2. Boxplots comparing performance of slaPRS (differing in choice of level 0 predictors from each block), *PRS*_*Marquez*_, and single population PRS: *PRS*_*EUR*_ & *PRS*_*AFR*_ (see methods) quantified through adjusted *R*^2^. Testing samples stratified by overall % of European ancestry.

While thus far we only considered the full slaPRS model 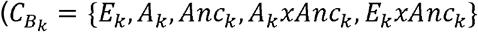,we then considered slaPRS under our alternative level 1 models that vary predictors from each local window. For the simplest case 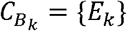 i.e. only European European GWAS considered and stacking local European PRS across blocks), slaPRS had adjusted *R*^2^ ranging from 3.28% for 0-20% European ancestry to 5.45% for 80-100% European Ancestry and noticeably outperformed *PRS*_*EUR*_ However, slaPRS under 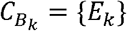 exhibited the strongest ancestry dependence (0.005 increase in adjusted *R*^2^ across ancestry quantiles) across all methods. For 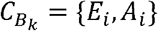 (i.e. integrating European and African GWAS and stacking local European and African PRS across blocks), slaPRS further increased performance (compared to the single population case 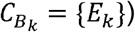 with adjusted *R*^2^ ranging from 5.77% to 6.27% and had noticeably reduced ancestry dependence (0.001 increase in adjusted *R*^2^ across ancestry quantiles). The full level 1 model 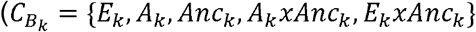 further added local ancestry with interaction terms and performed comparably to the previous model ignoring ancestry 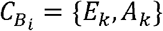. Negligible differences in the full model and the model excluding local ancestry were present only in simulations of complete sharing of transethnic genetic effects.

#### Effect of Overall Heritability, Number of Causal Variants, Window Size, and African GWAS Sample Size

We quantified how slaPRS fared against other approaches across different simulation settings including: overall heritability *h*^2^ ∈ {0.10,0.30}, number of causal variants *m* = {5,100,500,1000}, African GWAS sample size *n* ∈ {2000,5000,10000}, window sizes ∈{1*Mb*,5*Mb*} (see Supplementary), and training data size ∈ {3000,7000} (see Supplementary). Across all settings, slaPRS generally improved performance as compared to single ancestry PRS: *PRS*_*AFR*_ and *PRS*_*EUR*_ (Supplementary Figure 3). Two factors had a sizable impact on the performance of slaPRS generally and its comparison to *PRS*_*Marquez*_. The first major factor impacting PRS performance was the African GWAS sample size. As the African GWAS sample size decreased (while fixing *h*^2^ = 0.30, *m* = 100) the C+T *PRS*_*AFR*_ performed increasingly worse compared to other methods (Figure 3). The performance of the full slaPRS model similarly decreased as the African GWAS sample size decreased, reflecting less informative contributions about the true risk variants from the African cohort. Furthermore, slaPRS exhibited a stronger ancestry dependence (converging towards the European only slaPRS model) as the African GWAS sample size decreased: For every increase in European ancestry quantile, slaPRS under the full model had an average change in average adjusted *R*^2^of 0.0009, 0.001 and 0.003 for African GWAS sample sizes of n=10000, n=5000, and n=2000 respectively. However, even for the smallest African GWAS sample size scenario, slaPRS had the highest adjusted*R*^2^ across ancestry quantiles.

**Figure 3.**
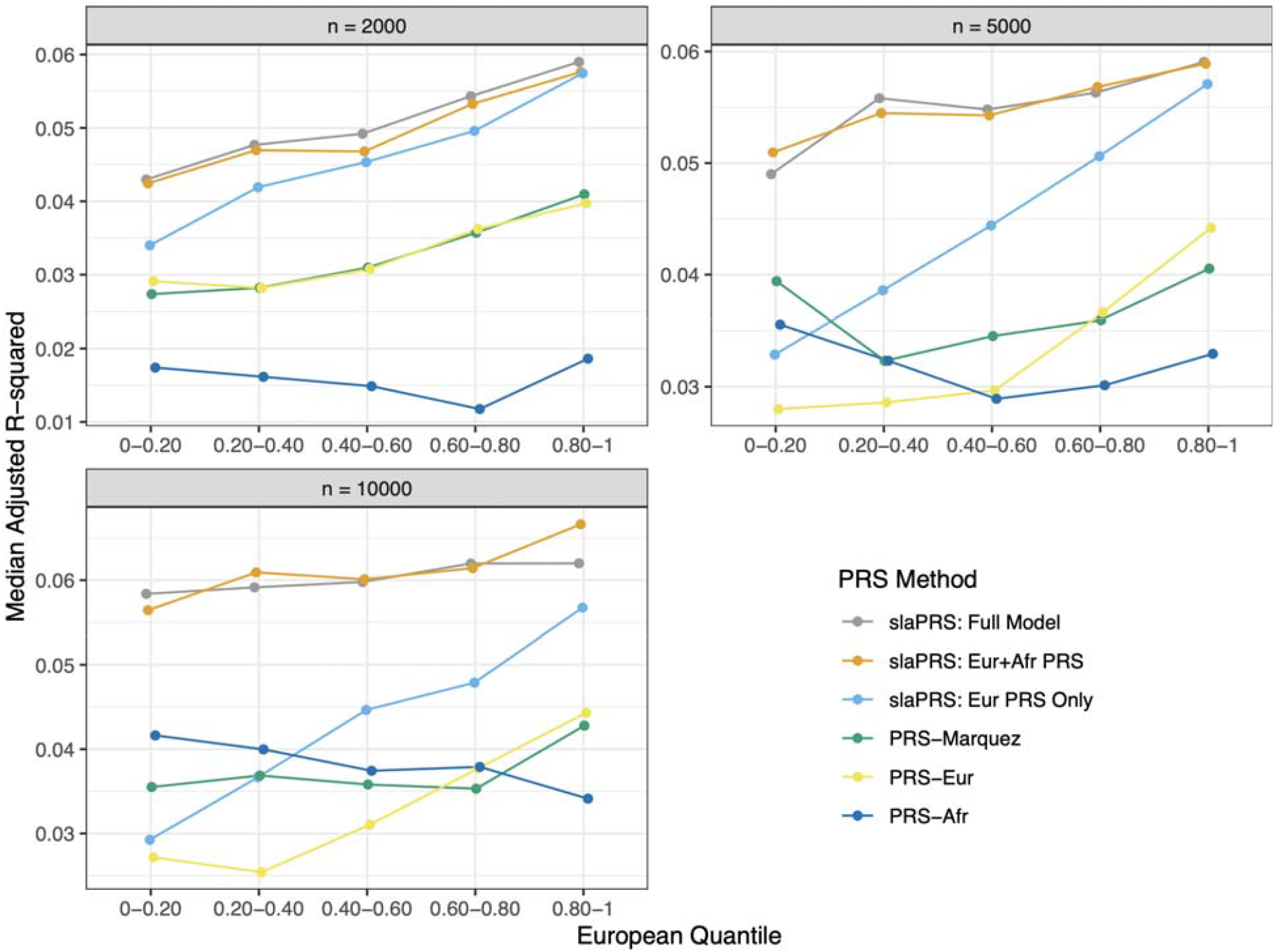
Line graph comparing PRS performance across methods (quantified by median adjusted *R*^2^ between estimated PRS and phenotype value) as the African GWAS sample size changes (n=2000, 5000, 10,000). Testing admixed samples stratified by European ancestry quantile.

The second factor impacting slaPRS, especially compared to *PRS*_*Marquez*_, was polygenicity and distribution of per variant effect sizes (Supplementary Figures 3, 4). slaPRS generally had the greatest improvement in polygenic (*m* = 100,500)simulations with moderate to large per variant effect sizes (*h*^2^ = 0.30,*m* = 100,500 and *h*^2^ = 0.10,*m* = 100) driving clear genetic signals. Under these simulation parameters, the median adjusted *R*^2^ of the full slaPRS model was 58.1% to 96.7% larger than the median adjusted *R*^2^of *PRS*_*Marquez*,_. In such settings, a majority of window’s local ancestry PRS contributing genetic signal to the stacking model. On the opposite end, when polygenicity was lower (*m* = 5 causal variants, *h*^2^ = 0.15)the median adjusted *R*^2^ for slaPRS was more similar to *PRS*_*Marquez*_ (23.4% increase), as a few large per variant effect sizes drive a small number of windows to dominate the genetic signal with remaining windows adding noise to the model. slaPRS similarly performed more similar to *PRS*_*Marquez*_ (21.1% and 27.3% increase in adjusted *R*^2^) in simulations of high polygenicity with low per variant effect sizes (*m* =500,1000 and *h*^2^ = 0.10), as most windows are uninformative and those with very small genetic signal are likely overly penalized and shrunk.

### 1.4.2 Comparison of PRS Performance Assuming Differences in Genetic Architecture across Ancestral Populations

We also considered simulations in which the genetic architecture differed across ancestral populations (i.e., unique population-specific effect sizes), causing population-specific GWAS to vary from both differences in LD and true underlying effects across populations. We computed slaPRS using GWAS effect sizes varying the transethnic genetic correlation across risk variants ρ = {0.2, 0.5, 0.8}. We again focused on our base simulation parameters (heritability *h*^2^ = 0.10, number of causal variants *m* =100, and equal size European and African GWAS sample size *n* =10,000). For the single population *PRS*_*EUR*_ and *PRS*_*AFR*_, which do not consider a risk variant’s local background, the adjusted *R*^2^ from the PRS model was stable in their corresponding admixed groups (80-100% European and 0-20% European) across changing transethnic genetic correlation. However, when transethnic genetic correlation was low (ρ = 0.2), *PRS*_*EUR*_ and *PRS*_*AFR*_ notably had an increased decay in PRS performance as the admixed ancestry group diverged from the population GWAS (Figure 4): Comparing the shared transethnic genetic architecture case vs when *ρ* = 0.20, the change in adjusted *R*^2^ was 0.005 vs 0.004 and -0.006 vs -0.001 across ancestry quantiles for *PRS*_*EUR*_and *PRS*_*AFR*_respectively. For slaPRS, notably the full level 1 stacking model 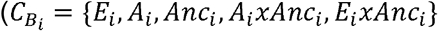 modeling local ancestry and interactions outperformed the model using only the local ancestry PRS 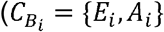 as the transethnic genetic correlation decreased. When genetic effects across ancestral populations were similar (*ρ* = 0.8)the percent increase in adjusted *R*^2^ between the full model and model ignoring local ancestry ranged from 10.9% to 14.3% across ancestry quantiles, as compared to 23.4% to 50.5% when transethnic genetic effects are vastly different (*ρ* = 0.2) (Figure 4). Notably, the overall adjusted *R*^2^ of the full level 1 model modeling ancestry specific effects dependent on a variant’s ancestral background was stable across values of *ρ* = {0.2,0.5,0.8}: (*R*^2^ = 5.27%,5.18%,5.67%) as compared to the model ignoring local ancestry (*R*^2^ =3.65%,4.09%,5.18%).

**Figure 4.**
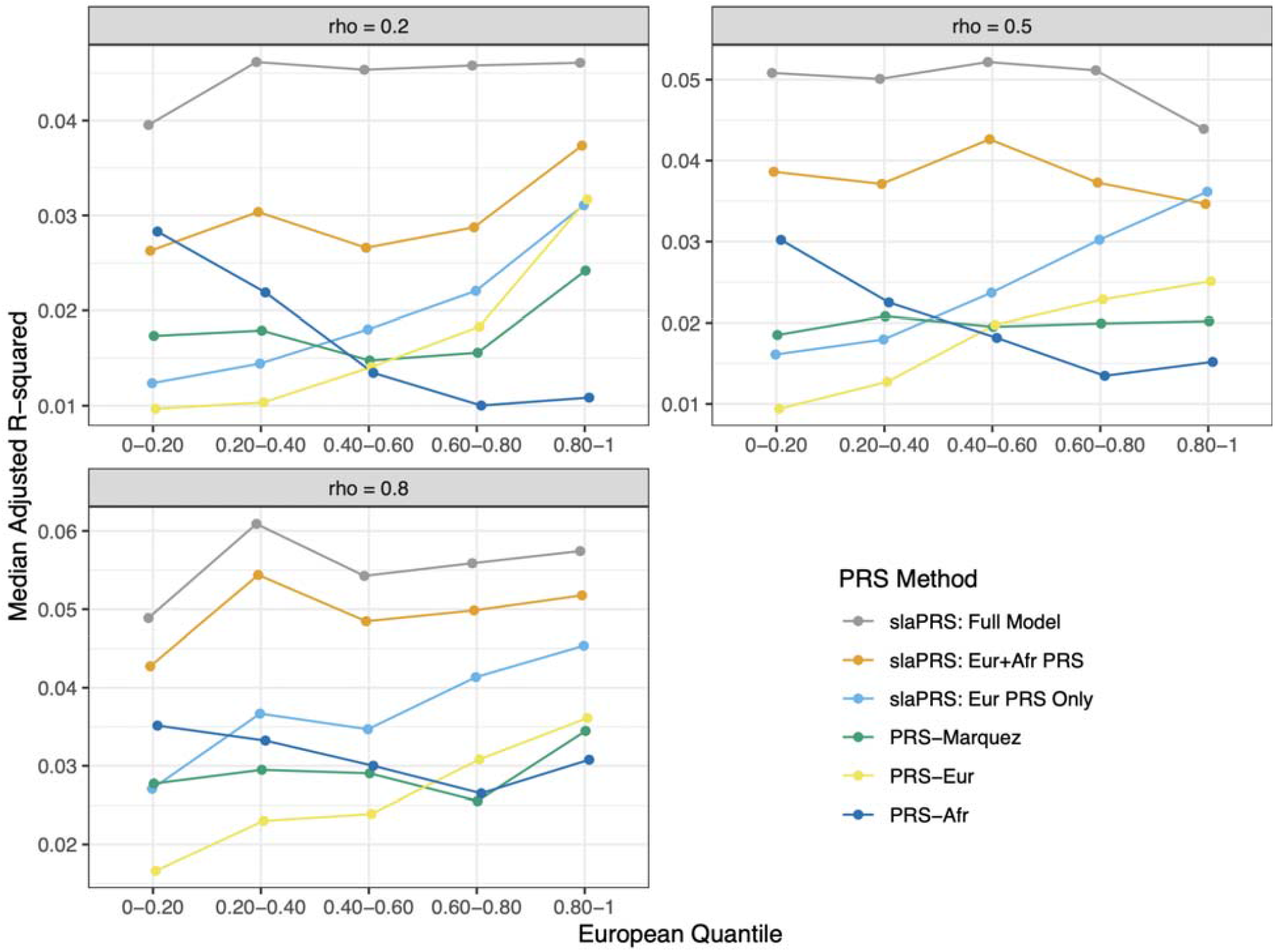
Line graph comparing PRS performance as quantified through median adjusted *R*^2^between the estimated PRS and phenotype value. Transethnic genetic correlation varies from *ρ* = {0.2,05,0.8} and testing admixed samples stratified by European ancestry quantile.

### 1.4.3 Real Data Application

We conducted a real data application of our stacking method slaPRS using genotype and phenotype data from the UK Biobank. We considered three quantitative lipid traits: HDL, LDL, and total cholesterol using estimated European and African American GWAS effect sizes from the Global Lipids Genetic Consortium (see methods for details). We first compared our approach to *PRS*_*EUR*_, *PRS*_*AFR*_ (C+T using European and African GWAS effect sizes separately), and *PRS*_*Marquez*_ (combining *PRS*_*EUR*_ and *PRS*_*AFR*_ globally) across all samples. For all three traits, slaPRS improved the median adjusted r squared values compared to *PRS*_*EUR*_ and *PRS*_*AFR*_ (Table 1). Similarly, slaPRS improved stratification ability as shown in larger mean phenotype values comparing individuals in the top and bottom 10% of the PRS distribution: HDL (0.373 vs 0.365, 0.324), LDL (1.019 vs 0.858, 0.905), TC (1.317 vs 1.028, 1.203). However, slaPRS performed similarly to *PRS*_*Marquez*_ across all three traits with respect to both metrics, a pattern observed in simulation scenarios of lower polygenicity causing fewer windows to contribute to trait heritability (Table 1). Across the three traits, only 1.6% (HDL), 6.6% (LDL), and 2.1% (TC) of all level 0 local population PRS across the genome had an *R*^2^ > 0.10 with the overall trait PRS. For LDL which had the highest signal to noise ratio, there was a minor improvement in *R*^2^both and top vs bottom 10% stratification ability for slaPRS. Furthermore, we found limited improvement in slaPRS using the full level 1 stacking model 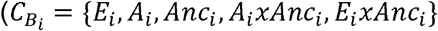 compared to the reduced model 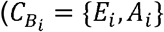

**Table 1.** Performance metrics for lipid phenotypes in UKB. a) Median adjusted *R*^2^from model PHENO ∼ PRS + PC1 + PC2 + PC3 + PC4. b) Difference in mean phenotype for individuals in top 10% of PRS distribution vs bottom 10%.

| Phenotype | Median Adjusted $R^2$ | | | | |
| --- | --- | --- | --- | --- | --- |
| | slaPRS<br>(Full Model) | slaPRS<br>(No Ancestry) | EUR C+T | AFR C+T | Global Stacked<br>($PRS_{Marquez}$) |
| HDL | 0.081 | 0.084 | 0.069 | 0.054 | 0.083 |
| LDL | 0.112 | 0.112 | 0.088 | 0.097 | 0.110 |
| TC | 0.115 | 0.113 | 0.093 | 0.091 | 0.112 |

Table 1. Performance metrics for lipid phenotypes in UKB. a) Median adjusted $R^2$ from model $PHENO \sim PRS + PC1 + PC2 + PC3 + PC4$ . b) Difference in mean phenotype for individuals in top 10% of PRS distribution vs bottom 10%.
|  | Mean Phenotype in Top vs Bottom 10% PRS Quantile |  |  |  |  |
| --- | --- | --- | --- | --- | --- |
| Phenotype | slaPRS<br>(Full Model) | slaPRS<br>(No Ancestry) | EUR C+T | AFR C+T | Global Stacked<br>($PRS_{Marquez}$) |
| HDL | 0.377 | 0.390 | 0.369 | 0.321 | 0.401 |
| LDL | 1.025 | 1.024 | 0.853 | 0.903 | 1.008 |
| TC | 1.317 | 1.307 | 1.039 | 1.202 | 1.330 |

We then stratified our testing samples by European ancestry quantile to 1) reassess overall PRS performance on admixed individuals in quantiles of 20%-80% European ancestry (removing primarily European or African admixed African British) and 2) quantify ancestry dependence of PRS performance across all five ancestry quantiles. In the bottom and top quantiles of predominantly homogenous African or European admixed African British, using single ancestry *PRS*_*EUR*_ and *PRS*_*AFR*_ tended to outperform. However, in the more heterogeneous admixed samples (20-80% European ancestry), slaPRS *PRS*_*Marquez*_ and had the best median adjusted across all methods with comparable results for the three traits: HDL (0.066 and 0.070), LDL (0.103 and 0.098), TC (0.079 and 0.081) (Figure 5). Regarding ancestry dependence of PRS method, across traits *PRS*_*EUR*_ and *PRS*_*AFR*_ exhibited the strongest ancestry dependence, performing better as the proportion of European or African ancestry increased. On the other hand, methods using multiple ancestry GWAS had reduced ancestry dependence, with slaPRS having the smallest dependence followed by *PRS*_*Marquez*_. For HDL, the average change in adjusted *R*^2^ for each European quantile increase for slaPRS, *PRS*_*EUR*_, *PRS*_*AFR*_, and *PRS*_*Marquez*_ was 0.004, 0.019, -0.006, and 0.011 respectively. LDL (−0.003, 0.014, -0.016, and 0.003) and TC (−0.002, 0.012, -0.014, -0.005) had similar patterns across methods.

**Figure 5.**
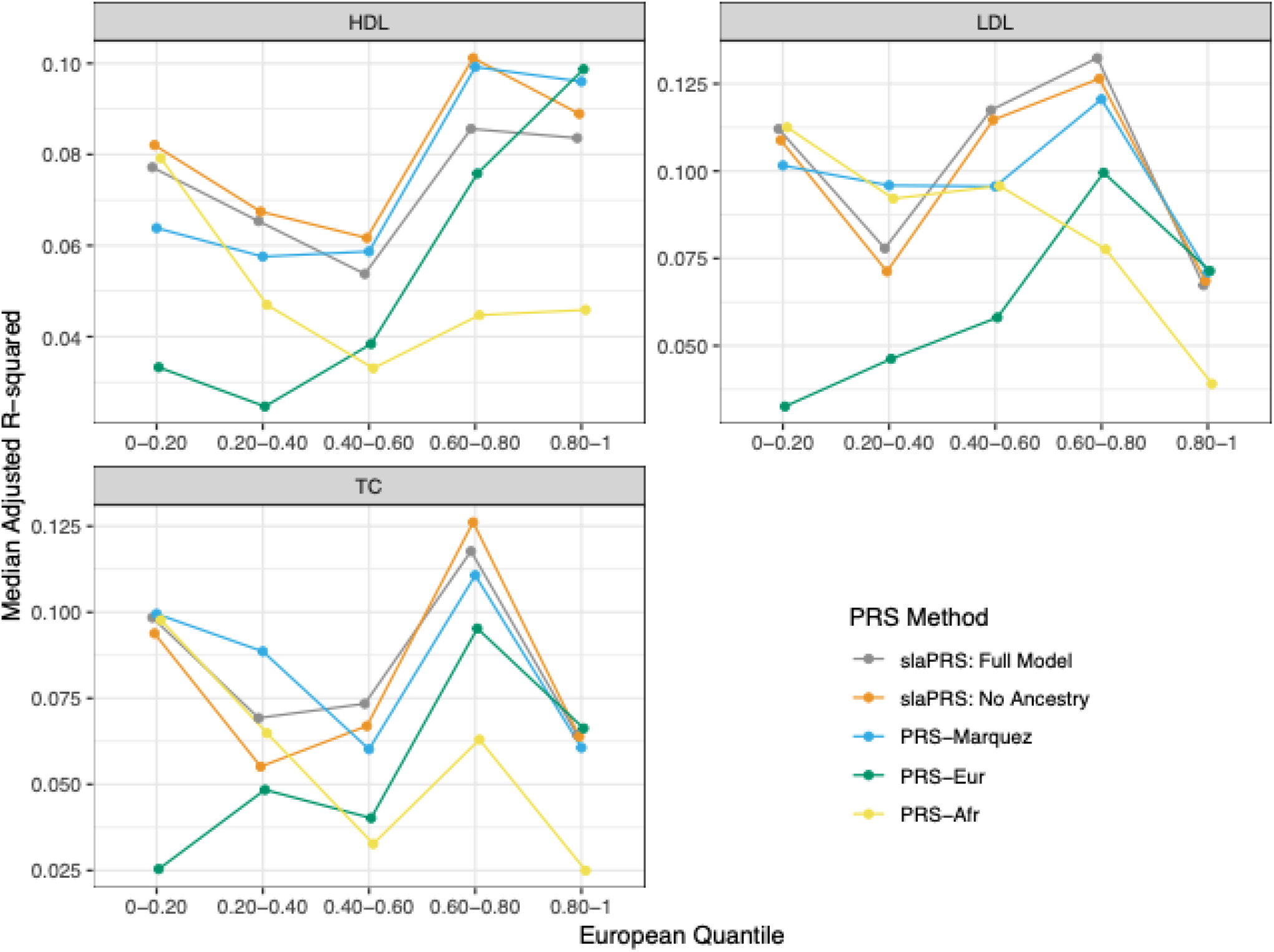
Line graph comparing PRS Performance for UKB lipid phenotypes. Performance quantified through median adjusted *R*^2^ from model PHENO ∼ PRS + PC1 + PC2 + PC3 + PC4. Testing admixed samples are stratified by European ancestry quantile.

## 1.5 Discussion

In this work we proposed a novel stacking framework to locally incorporate GWAS from multiple populations into construction of PRS for admixed individuals. Our method, slaPRS, segments admixed genomes into local regions of varying ancestry and optimizes a linear combination of local population specific PRS, local ancestry, and potential interactions. In simulations, we first recapitulated previous findings that traditional PRS constructed using a single population GWAS in admixed samples are ancestry dependent. We then showed across a range of genetic architectures (varying heritability, number of causal variants, underrepresented GWAS sample ancestry size, and transethnic genetic correlation across ancestral populations) that slaPRS can outperform existing approaches (*PRS*_*EUR*_, *PRS*_*AFR*_ and *PRS*_*Marquez*_) and reduce the dependence compared to *PRS*_*EUR*_ and *PRS*_*AFR*_. In real data, we leveraged ancestry specific GWAS for lipid traits from the Global Lipids Genetic Consortium to compare slaPRS to existing PRS methods in admixed African British from the UK Biobank. We found in these lipid traits that incorporating multiple ancestry GWAS similarly improved performance and strongly reduced the ancestry dependence of PRS performance.

From our simulations and real data applications, we conclude that slaPRS for PRS in admixed individuals is likely optimal (compared to existing approaches) for traits with high heritability and polygenicity. slaPRS extends *PRS*_*Marquez*_ to combine information locally as opposed to globally and comparisons had interesting findings. In simulations, we found the smallest improvements were in trait architectures with low polygenicity (few windows meaningfully contribute to trait heritability with others add noise to the model) or in highly polygenic settings where per-variant effect sizes are small (hard to distinguish signal from noise and genetic signals may be over shrunk). In real data applications, we found slaPRS and *PRS*_*Marquez*_ performed similarly across the three lipid traits, likely driven by their trait genetic architecture. For the lipid traits studied, the former simulation scenario may be most prevalent as only 2-6% of all local PRS across windows contributed to the estimated PRS causing most regions to solely add noise to the model. As a result, noticeable improvements in slaPRS over *PRS*_*Marquez*_ may be observed in more heritable and polygenic traits, such as height, in which more local windows across the genome will contribute genetic signal.

However, evaluating slaPRS had a surprising finding that explicitly modeling local ancestry in the slaPRS model (vs the model excluding local ancestry) had the most improvement when there existed at least moderate heterogeneity in true causal variant effect sizes across ancestral backgrounds. In simulations, this was shown through the largest increase in PRS performance between slaPRS models when transethnic genetic correlation was low (*ρ* = 0.20), with no improvements under scenarios of shared transethnic genetic architecture. In lipid traits from the UK Biobank, we observed similar findings regarding modeling local ancestry. In such traits, modeling local ancestry in the slaPRS model only provided marginal improvements, consistent with high estimated transethnic genetic correlations from Million Veteran Program participants for HDL (*ρ* = 0.84) and moderate correlation for the other traits (*ρ* ∈ [0.47,0.69])^41^. High transethnic genetic correlations for the considered lipid traits are consistent with recent findings from Hou et al, that suggest a majority of common traits likely have similar causal effects across populations^18^. Such findings have immediate implications, as slaPRS and other approaches considering local ancestry background may find the most improvement in traits with significant differences in transethnic genetic architecture.

Historically in genetic studies, individuals are often discretized into ancestral populations and treated as homogenous within the group. Ding et al has recently challenged the historical paradigm by showing PRS accuracy varies between individuals even within a “homogenous” genetic ancestry cluster to ultimately push for treating genetic ancestry on a continuum^22^. Our method slaPRS is tailored to treat genetic ancestry on a continuum by taking a local approach to PRS prediction in admixed samples. As mentioned, *PRS*_*Marquez*_ previously combined global population specific PRS successfully in admixed individuals, though in doing so uses a single weight for population specific effects. Potential heterogeneity in true population specific risk variants, estimated population specific GWAS effect sizes, and admixture proportions across loci and individuals would cause use of a single weight to be suboptimal. slaPRS extends *PRS*_*Marquez*_ by combining population specific PRS at the local level instead to 1) allow for varying effects of local population specific PRS across the genome and 2) increase overall external GWAS sample sizes to improve effect size estimation and identify the true causal variants. The first benefit is accomplished through our level 1 elastic net stacking model that learns a linear combination of local population specific PRS (and local ancestry with interaction effects) to inform which population’s local PRS should be upweighted or shrunk. In the case that the true causal effect differs due to ancestral background, slaPRS handles this scenario by modeling the local ancestry and interactions with the local population specific PRS, allowing for the effect of a local population specific PRS to differ based on its ancestral background. The second benefit is accomplished by increasing the overall effective GWAS sample size through incorporating information from each population’s GWAS. In the case that the genetic architecture is shared across ancestral backgrounds, using information from both GWAS will boost power and improve effect size estimation of the shared risk variants and their locations. However, when the genetic architecture differs across populations it is unclear whether using multiple population GWAS can be viewed in a similar manner.

slaPRS has desirable statistical and computational properties as well. First, similar to other machine learning-based PRS methods such as TL-PRS^44^ in the context of cross population prediction incorporating multiple ancestry GWAS, slaPRS avoids the needs for any distributional assumptions on transethnic effect sizes as compared to the cross population PRS methods PRS-CSx^21^ and PolyPred^45^ (Utilizes BOLT-LMM^46^ and PRS-CS^47^ which treat SNP effects as random). As a result, our approach makes no assumption on whether a risk variant is shared across population, where each local population PRS in a genomic region can include its own set of risk variants. Second, slaPRS does not require an external LD reference panel or genotypes outside of the admixed genotypes. Third, slaPRS can accommodate any PRS algorithm to construct local population PRS (here we use the C+T algorithm for simplicity). For example, REGENIE^32^ uses a ridge regression based approach to construct level 0 local PRS before stacking. Lastly, our approach is computationally very efficient, as discretizing the genome into local windows facilitates efficient parallel processing of level 0 predictions, with a final level 1 elastic net model that can be fit very fast with standard statistical packages.

While slaPRS provides a novel stacking approach to combine population specific GWAS information locally, it has a few limitations to consider. We assume existence of GWAS from each ancestry contributing to a genetic admixture, though high powered GWAS in understudied homogenous populations such as Africans are currently limited or non-existent. As a result, our real data application was limited to using African American GWAS as proxies for African GWAS, with only a handful of lipid traits from the Global Lipids Genetic Consortium having sufficiently large GWAS sample sizes. Recent efforts for genomic research in diverse populations such as the African biobank^48^ should help to resolve this issue. Furthermore, we describe our framework for continuous value phenotypes, owing to currently limited access to large sample GWAS for binary case/control traits in each ancestral population. Extending this framework to case/control traits using a logistic regression elastic net and liability threshold model should be straightforward. Lastly, while we push to treat admixed individuals on a genetic ancestry continuum, our approach assumes the super population groups such as “European” and “African” have homogenous genetic architecture with respect to a complex trait across their subpopulations. However, studies have shown a high degree of genetic diversity across the African continent^49,50^ with unique demographic histories driving substantial cultural and ethnic differences that may cause treating all African subpopulations as homogenous to be problematic^22,51^.

Despite the limitations, slaPRS provides an efficient data driven framework to constructing polygenic risk scores in admixed samples that leverage multiple population GWAS. In providing a method that not only performs well in admixed samples, but equally well across varying ancestry proportions we strive to improve on the current inequity in genetics research that is fast resolving in our community. Furthermore, as sample sizes increase in underrepresented populations for more traits, we expect slaPRS to have additional applications. Lastly, while our work thus far only considered two-way admixture, our approach can easily accommodate three or more ancestral populations and respective external GWAS. In coming years admixture will likely extend beyond the historically predominant African American and Latino admixed groups as people and cultures from various ancestral backgrounds are brought together geographically. As a result, we believe our method’s flexibility to accommodate increasingly complex admixture types using information from multiple GWAS will become even more relevant.

## Data Availability

This research used genetic and phenotypic data from the UK Biobank Resource under Application Number 24460. Data is available for download for approved researchers of the UK Biobank. High powered ancestry specific GWAS from the Global Lipids Genetics Consortium are publicly available: http://csg.sph.umich.edu/willer/public/glgc-lipids2021/.

## Code Availability

slaPRS and necessary functions has been implemented as an R package and can be installed via running devtools::install_github(‘kliao12/slaPRS’) using the devtools library in R. An example workflow is available at https://github.com/kliao12/slaPRS

## Acknowledgements

Funding for this project was provided by National Institutes of Health grant R01 HG011031 and R01 HG005855 (S.Z.) and the NIH/National Human Genome Research Institute Genome Science Training Program (T32HG00040). We thank UK biobank participants and study teams for providing high quality genetic and phenotypic data. Lastly, we appreciate the helpful insights and feedback given by Dr. Jean Morrison on general method development and analysis.

## 1.6 Supplementary

### 1.6.1 Derivation of weighted function learned from slaPRS

We restate our model setup consisting of a sample of N admixed individuals with ancestral contributions from population A and B. Let **X** be the *NxM* admixed genotype matrix (M is the total number of variants genome wide) and **Y** the *Nx*1phenotype vector. Let *L*_ij_ be an *NxM* matrix denoting the haplotype-level local ancestry (*l*_ijl_, *l*_*ij2*_.) of individual *I* at marker *j*. We assume the phenotype can be expressed as:

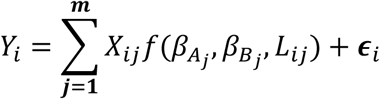

Where *X*_ij_ is the genotype dosage for individual *I* at marker *j* and 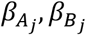 are effects for marker *j* on the phenotype in populations A and B respectively. Here, 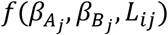 is a weighted average of population specific GWAS effect sizes and local ancestry learned via our stacking approach.

Following construction of level 0 model predictions in each window 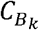 across the genome (includes local population A PRS *A*_*k*_ and local population B PRS *B*_*k*_, local ancestry, and interaction terms) we fit the following stacking model:

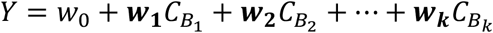

Expanding out terms for the k-th window:

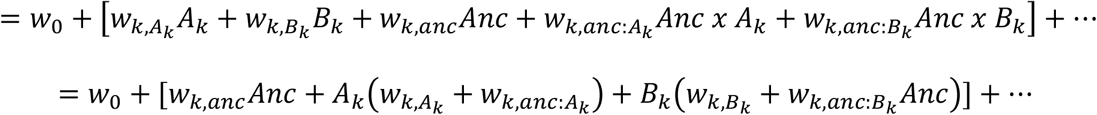

The stacking procedure learns a linear combination of level 0 model prediction in each window 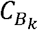 across the genome through estimating the weights *w*_*k*_.*A*_*k*_ and *B*_*k*_ are themselves weighted sum of risk variants using population specific GWAS reducing the form to:

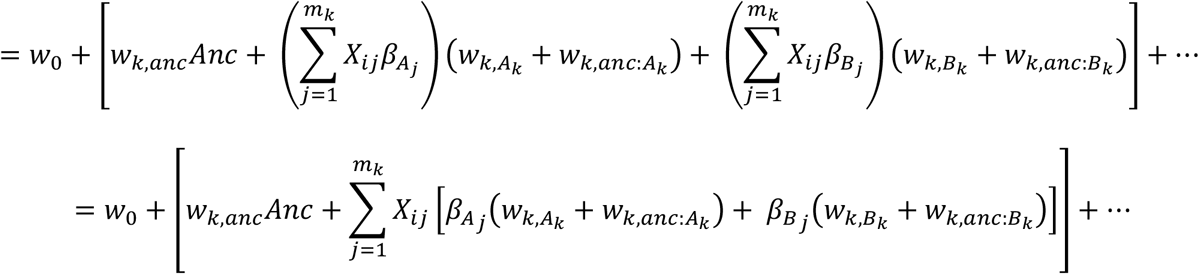

Where 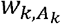 and 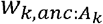 are weights for population A specific local PRS *A*_*k*_ and its local ancestry interaction term. Because *A*_*k*_ (and likewise for *B*_*k*_ is a function of population A GWAS effect sizes that is shared across all variants in the window *k*, we replace the notation 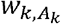 with 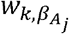 and similarly *anc*_*k*_ is a function of *L*_*ij*_ so we replace 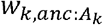 with 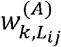.

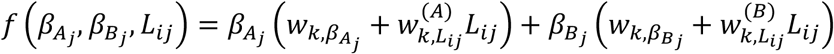

### 1.6.2 Effect of window size and training dataset size

slaPRS takes a sliding local window approach to construct local population-specific polygenic risk scores and thus may be sensitive to the size of the window. In simulations under our base scenario (*h*^2^ = 0.10, *m* = 100)we considered both 1Mb and 5Mb windows. PRS performance quantified by adjusted *R*^2^with the phenotype were highly consistent across window sizes suggesting slaPRS is robust to window size (Supplementary Figure 5). We further quantified the effect of varying the training dataset size of admixed individuals (n = 3000, n = 7000). As compared to *PRS*_*EUR*_ and *PRS*_*AFR*_., slaPRS uses the training data to weight local population specific PRS (and the variants effects themselves) and increased performance should be dependent on the training dataset size. In general, slaPRS for training sizes n=3000 and n=7000 generally had increased adjusted *R*^2^when the training size was larger compared to *PRS*_*EUR*_ (77.3%, 84.9%), *PRS*_*AFR*_ (35.3%,66.1%) and *PRS*_*Marquez*_ (64.4%,66.4%)

### 1.6.3 Supplementary Tables and Figures

**S Figure 1.**
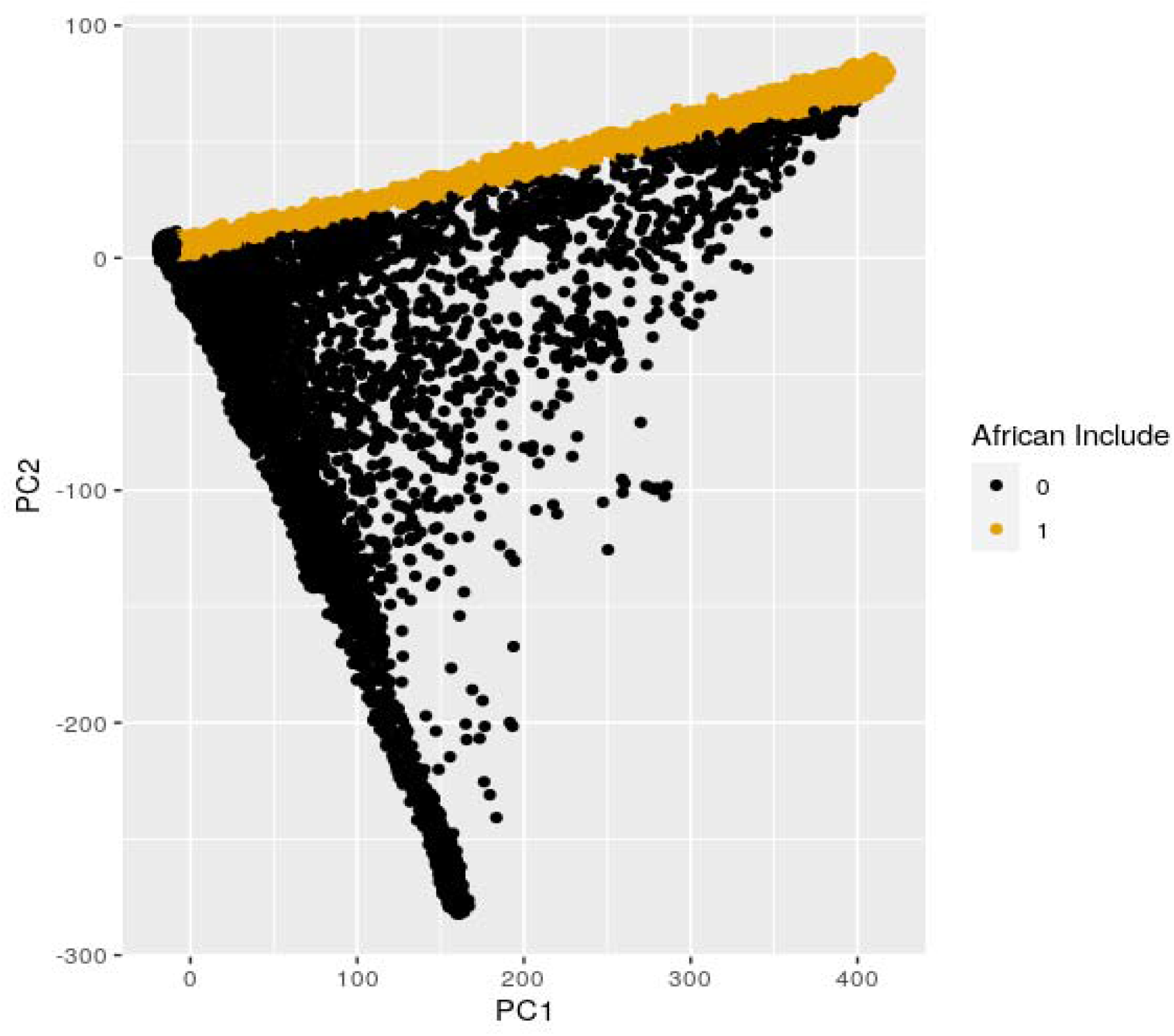
Scatterplot of n=20,262 UKB samples containing African ancestry along diagonal of PC1.

**S Figure 2.**
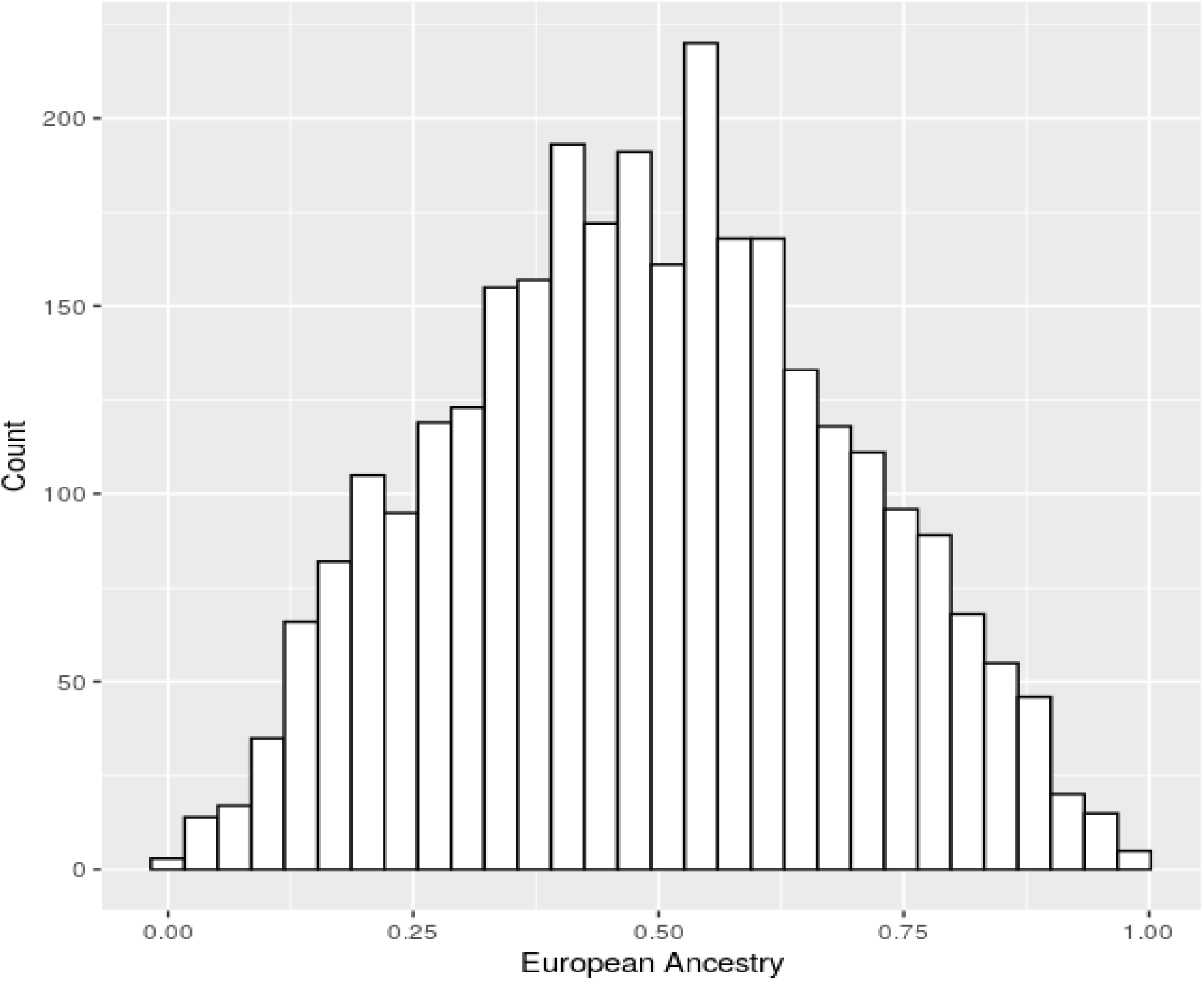
Histogram of the distribution of overall European ancestry across n=10,000 simulated admixed African Americans (for a single simulation).

**S Figure 3.**
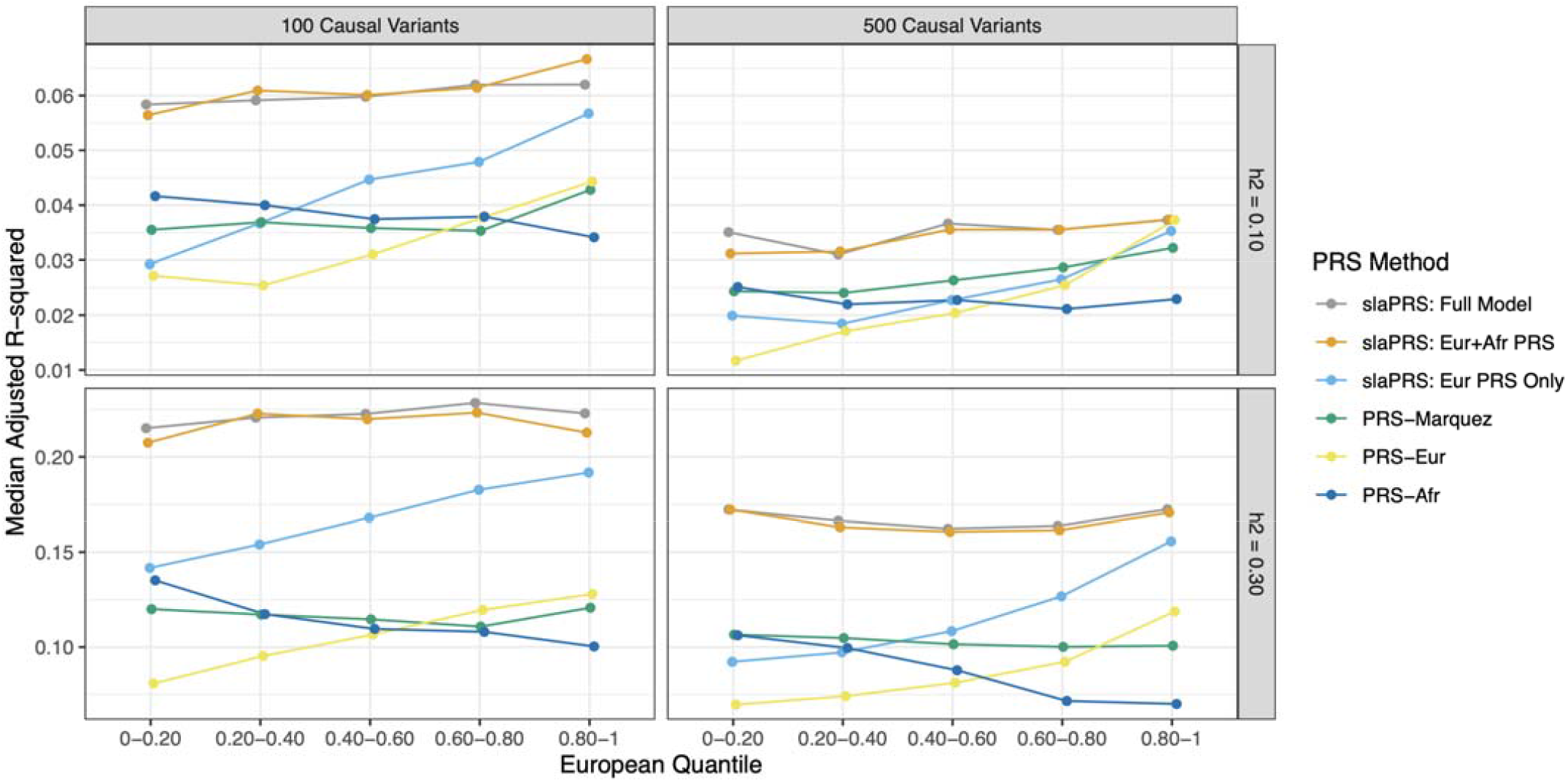
Line graph comparing PRS performance across PRS methods for different simulation settings using adjusted *R*^2^ between estimated PRS and simulated phenotype. Simulation parameters: heritability (*h*^2^=0.1,0.3) and number of causal variants (m=100,500)

**S Figure 4.**
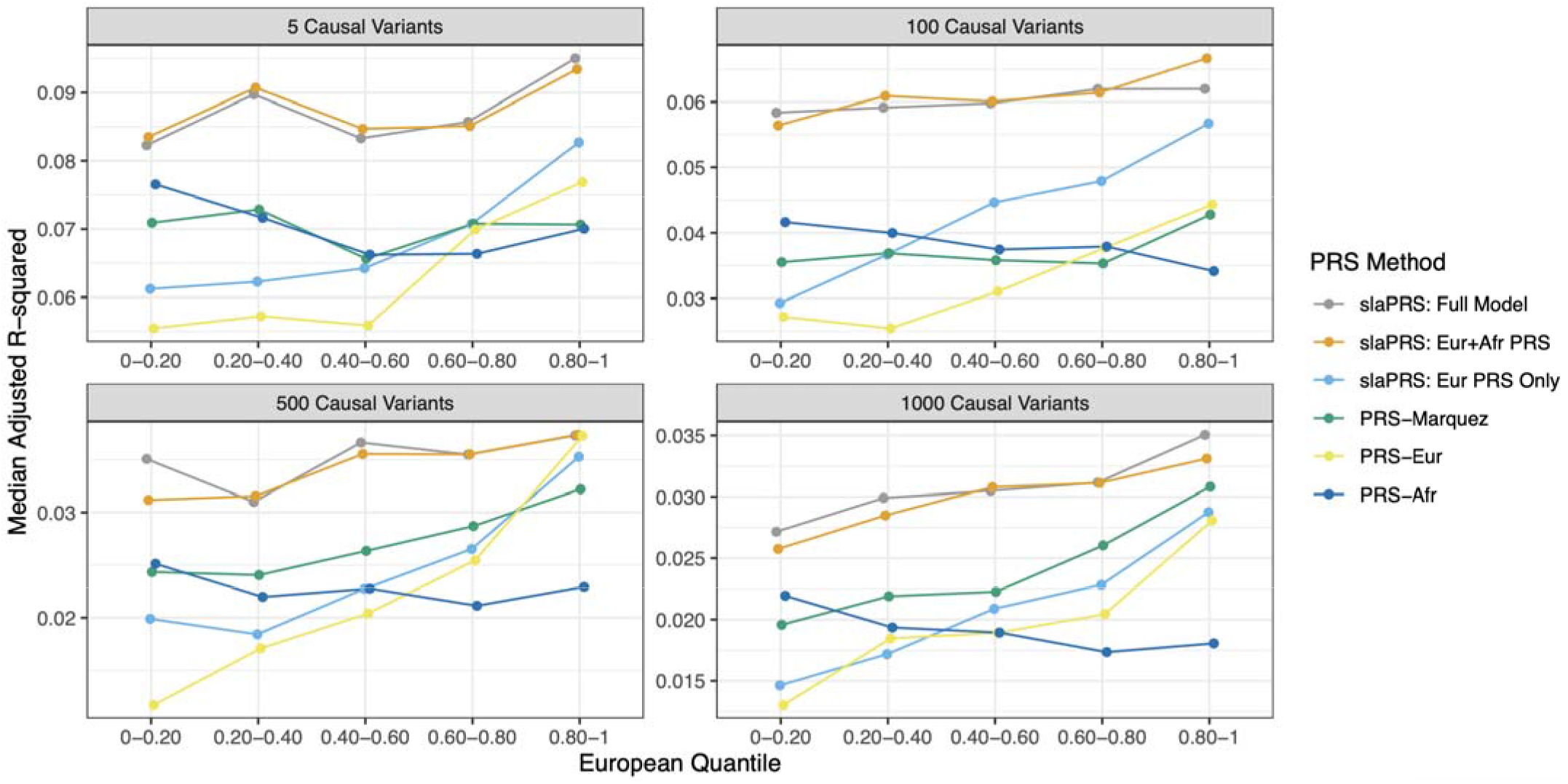
Line graph comparing PRS performance across PRS methods for different simulation settings using adjusted *R*^2^between estimated PRS and simulated phenotype. Simulation parameters: heritability (*h*^2^=0.1) and number of causal variants (m=5,100,500,1000).

**S Figure 5.**
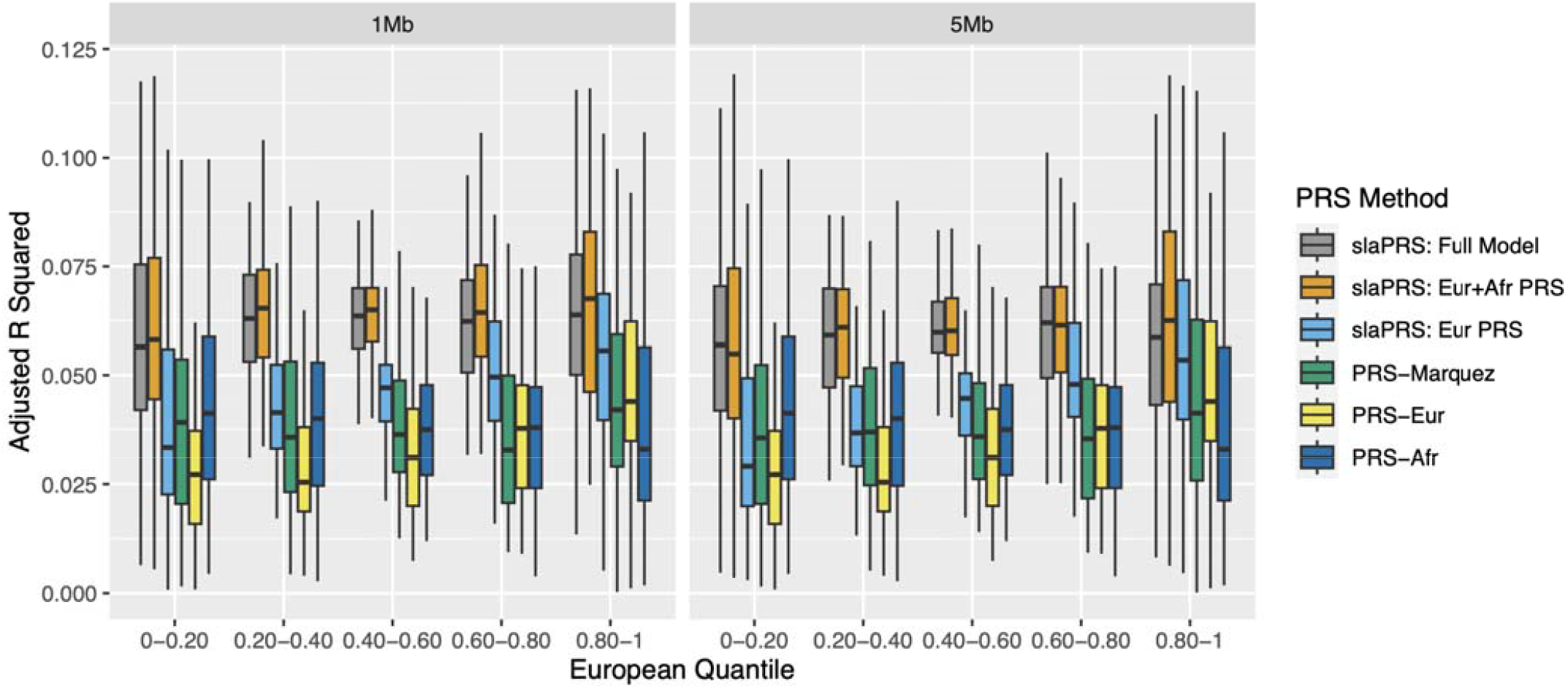
Comparison of PRS performance across methods (quantified by adjusted between estimated PRS and phenotype value) as the window size in slaPRS varies (1Mb, 5Mb).

